# Understanding the nature of face processing in early autism: A prospective study

**DOI:** 10.1101/2020.05.06.20092619

**Authors:** Charlotte Tye, Giorgia Bussu, Teodora Gliga, Mayada Elsabbagh, Greg Pasco, Kristinn Johnsen, Tony Charman, Emily J.H. Jones, Jan Buitelaar, Mark H. Johnson, the BASIS team

## Abstract

Dimensional approaches to psychopathology interrogate the core neurocognitive domains interacting at the individual level to shape diagnostic symptoms. Embedding this approach in prospective longitudinal studies could transform our understanding of the mechanisms underlying neurodevelopmental disorders. Such designs require us to move beyond traditional group comparisons and determine which domain-specific atypicalities apply at the level of the individual, and whether they vary across distinct phenotypic subgroups. As a proof of principle, this study examines how the domain of face processing contributes to a clinical diagnosis of Autism Spectrum Disorder (ASD). We used an event-related potentials (ERPs) task in a cohort of 8-month-old infants with (n=148) and without (n=68) an older sibling with ASD, and combined traditional case-control comparisons with machine-learning techniques like supervised classification for prediction of clinical outcome at 36 months and Bayesian hierarchical clustering for stratification into subgroups. Our findings converge to indicate that a broad profile of alterations in the time-course of neural processing of faces is an early predictor of later ASD diagnosis. Furthermore, we identified two brain response-defined subgroups in ASD that showed distinct alterations in different aspects of face processing compared to siblings without ASD diagnosis, suggesting that individual differences between infants contribute to the diffuse pattern of alterations predictive of ASD in the first year of life. This study shows that moving from group-level comparisons to pattern recognition and stratification can help to understand and reduce heterogeneity in clinical cohorts, and improve our understanding of the mechanisms that lead to later neurodevelopmental outcomes.

**General Scientific Summary:** This study suggests that neural processing of faces is diffusely atypical in Autism Spectrum Disorder, and that it represents a strong candidate predictor of outcome at an individual level in the first year of life.

## Introduction

Autism spectrum disorder (ASD) is defined on the basis of social and communication impairment, restricted patterns of behaviours and interests, and sensory anomalies in early childhood (American Psychiatric Association, 2013). ASD is characterised by high heterogeneity, expressed as considerable variability across individuals in terms of both clinical manifestations and underlying biology (E. J. Jones, Gliga, Bedford, Charman, & Johnson, 2014; Lai, Lombardo, Chakrabarti, & Baron-Cohen, 2013; Vorstman et al., 2017). Parsing this heterogeneity is a main theme of theoretical initiatives in mental health research, such as the Research Domain Criteria (RDoC) Framework (Insel et al., 2010). Theoretical models like RDoC propose a shift from unitary diagnostic labels towards determining how underlying impairments in a set of core domains contribute to diagnostic phenotypes. This may facilitate greater individualization of treatment options by enabling individuals to be characterized by dimensional scores reflecting domain functioning, hypothetically facilitating neurobiologically-informed treatments. Studying RDoC domains in early development, prior to the onset of behavioural symptoms, might be particularly critical for understanding how alterations in these domains contribute to symptom emergence. To do this, it is fundamental to adopt analytic strategies that profile a selected domain at the individual level, and understand its developmental link to ASD outcome.

While most genetic studies treat ASD as a unitary clinical category, the majority of ASD risk is thought to be spread across many genes with individually small and pleiotropic effects (Huguet, Benabou, & Bourgeron, 2016). A recently proposed framework suggests that these genetic factors act through the critical aggregation of earlier-interacting liabilities in contributing to the later clinical expression of ASD (Constantino, 2018). These liabilities are best described as endophenotypes, quantitative heritable neuropsychiatric alterations that can be identified in the general population as continuously distributed traits (Bearden & Freimer, 2006). A leading candidate domain in the mechanisms underlying ASD development is social cognition, and more specifically, face processing (G. Dawson, Bernier, & Ring, 2012; G. Dawson et al., 2005). Altered face processing has been shown to be a strong candidate precursor of ASD diagnosis and its social features (Mayada Elsabbagh et al., 2012; E. Jones et al., 2016), and represents one of the early measurable liabilities that could be aggregated with other factors to ultimately lead to ASD. However, little is known about *how and what* atypicalities in face processing contribute to a later ASD outcome.

From the first year of life, infants with later ASD demonstrate emerging atypicalities in social-communicative behaviour, such as a declining interest in human faces (W. Jones & Klin, 2013; Maestro et al., 2002; Osterling & Dawson, 1994). These behavioural changes appear to be accompanied by atypical neural responses to faces as measured by event-related potentials (ERPs), which provide the resolution required to investigate different temporal stages of information processing and can be obtained at younger ages than behavioural assessments (de Haan, 2007). While some studies have suggested that low-level sensory sensitivity to faces in infancy is associated with better social development (E. Jones, Dawson, & Webb, 2018), atypicalities in higher-level cortical processing of faces and gaze have been reported in toddlers and children with ASD (Geraldine Dawson et al., 2002; Grice et al., 2005; Sara Jane Webb et al., 2011). Altered responses to faces versus non-faces, and reduced differentiation of faces that shift gaze towards versus away from the viewer from 6 months of age are observed in infants with later ASD, as indexed by higher-level cortical responses (Mayada Elsabbagh et al., 2012; E. Jones et al., 2016). In ‘social first’ theories, atypicalities in social engagement and information processing mutually amplify each other over developmental time, reducing opportunities for social learning and contributing to the atypical development of social communication that is characteristic of ASD (M. Elsabbagh, 2020). Thus, examining neural responses to faces in infants with later ASD provides an excellent context in which to interrogate the generalisability or specificity of the mechanisms through which socio-cognitive atypicalities relate to ASD emergence.

Whilst previous research converges in identifying face processing as a relevant domain in ASD, studies have report greater divergence in the *nature* of ASD-associated atypicalities. Neural responses to faces can be characterized by many different features of an averaged waveform, which are hypothesized to reflect different underlying cognitive processes. These ERP components are typically identified by their polarity and timing. Although the general conclusion is one of altered face processing, this diversity could reflect a) consistent differences in lots of different specific aspects of face processing that are masked in different studies by their theory-driven focus on one or two components; b) individually-specific profiles of specific atypicality that will or will not be apparent at the group level depending on its composition; and/or c) distinct profiles of atypicality in particular coherent subgroups of individuals with ASD, again apparent or not at the group level depending on the sample composition. To understand these patterns, we need to couple both top-down and data driven analytic strategies with larger samples and individual-level data analysis.

Here, we used three analytic strategies to ask how atypicalities in the domain of face processing contribute to later ASD. First, we used a prospective approach because it enables the investigation of causal mechanisms. Based on a sibling recurrence rate of around 20% (Ozonoff et al., 2011), research on infants with an older sibling with ASD (“infant siblings”) has accumulated over the past decade, with concomitant progress in characterising candidate neural and cognitive precursors of symptom emergence in ASD (E. J. Jones et al., 2014; Szatmari et al., 2016). Prospective studies of infant siblings represent a powerful research design to identify such precursors (E. J. H. Jones et al., 2019), and hold the potential to generate developmental data that could transform our understanding of the nature of the ASD diagnostic category itself. Second, we combine group-based comparisons with investigating individual effects through a data-driven multi-feature machine learning approach. Since domains are multifaceted rather than single cognitive processes, such approach enabled us to examine the consistency of our results across both top-down prediction and bottom-up discovery and build a robust multivariate model for data integration and prediction of later clinical outcome at an individual level (Arbabshirani, Plis, Sui, & Calhoun, 2017; Rosenberg, Casey, & Holmes, 2018; Yahata, Kasai, & Kawato, 2017). Third, we decomposed heterogeneity through stratifying the ASD group with a clustering approach based on the domain under investigation (here social cognition/face processing) to examine whether subgroups showed qualitatively different face processing atypicalities (Lombardo et al., 2016; Zhao & Castellanos, 2016). Moving from group-level comparisons, to pattern recognition and stratification, this study promotes the use of novel, individual-level approaches for a dimensional understanding of brain development.

## Methods and Materials

### Participants

This study included 247 infants at elevated likelihood (EL), based on having an older biological sibling with ASD, and at typical likelihood (TL) of developing ASD, recruited from the British Autism Study of Infant Siblings (www.basisnetwork.org) across two independent cohorts. Specifically, 54 EL (21 male) and 50 TL infants (21 male) participated in cohort 1 (Mayada Elsabbagh et al., 2012), and 116 EL (64 male) and 27 TL (14 male) in cohort 2. TL controls were full-term infants (gestational age 38-42 weeks) recruited from a volunteer database at the Birkbeck Centre for Brain and Cognitive Development. Infants were seen for the face/gaze ERP task when they were approximately 8 months old *(Table S1/S2)*. Subsequently, 241 were seen for assessment around their third birthday by an independent team. Two TL children were absent for the 36-month visit but were included in the analysis as they showed typical development at the previous visits. Among the remaining 245 infants, 29 were excluded based on quality of EEG data, resulting in a final sample of 216 infants (TL=68, EL-no ASD=115, EL-ASD=33; see *Table S3* for details). All procedures were in agreement with ethical approval granted by the London Central NREC (approval codes 06/MRE02/73, 08/H0718/76), and one or both parents gave informed consent to participate in the study.

### Clinical assessment

The Autism Diagnostic Observation Schedule (ADOS)-generic(Lord et al., 2000), a semi-structured observational assessment, and the Social Communication Questionnaire (SCQ(Rutter, 2003)), a screening tool for ASD, were used to assess current symptoms of ASD at 36 months. The Autism Diagnostic Interview – Revised (ADI-R), a structured parent interview, was completed with parents of EL infants in cohort 1 and all children in cohort 2. These assessments were conducted without blindness to risk-group status by (or under the close supervision of) clinical researchers with demonstrated research-level reliability. The Mullen Scales of Early Learning (MSEL;Mullen, 1995) and the Vineland Adaptive Behavior Scale [VABS] (Sparrow, Balla, & Cicchetti, 1984) were used to measure, respectively, cognitive abilities and adaptive functioning at each visit.

Experienced researchers determined the best estimate clinical outcome by reviewing all available information from visits performed. Of the 148 EL participants included in analyses, 33 [22.3%] participants met criteria for ASD (hereafter EL-ASD) and the remaining 115 [77.7%] participants did not meet criteria for ASD (hereafter EL-no ASD), using ICD-10 criteria (cohort 1) or DSM-5 (cohort 2). There was a significant difference in clinical outcome by sex (*χ*^2^(2) = 13.5, p = 0.001), with more males receiving an ASD diagnosis than females (odds ratio, OR = 4.84; 95% confidence interval [CI; 1.93 to 12.1]; p<0.001).

### Electrophysiological measures

The task was the same as in Elsabbagh et al. (2012). It was designed to assess responses to the following contrasts: (1) faces (valid static (irrespective of gaze direction) vs. visual noise stimuli presented at the beginning of each block); (2) static gaze (faces with direct vs. averted gaze); and (3) dynamic gaze shifts (gaze toward vs. away from the infant). Components P100, N290, and P400 averaged across occipito-temporal channels were quantified by amplitude and latency in response to the different stimuli and the specified contrasts, and used as input features for subsequent analyses. See *Supplemental Materials* for details.

### Statistical analysis

#### Group-based comparison

A repeated measures ANOVA was conducted on each ERP parameter, with contrast as the within-subjects factor and group as the between-subjects factor (*Figure 1.A*). A set of analyses was run with cohort as an additional between-subjects factor and followed up with post-hoc t-tests to compare ERP amplitude and latency of the EL-ASD group against other groups. Sidak correction was used to correct for multiple testing. Covariates (age at time of EEG acquisition, MSEL visual reception and fine motor (non-verbal) t-score at 36 months) were entered into a second round of analyses. Gender was not a significant covariate in any analysis and was not retained. Analyses were performed on SPSS v22 (http://www.ibm.com/analytics/us/en/technology/spss).

**Figure 1:**
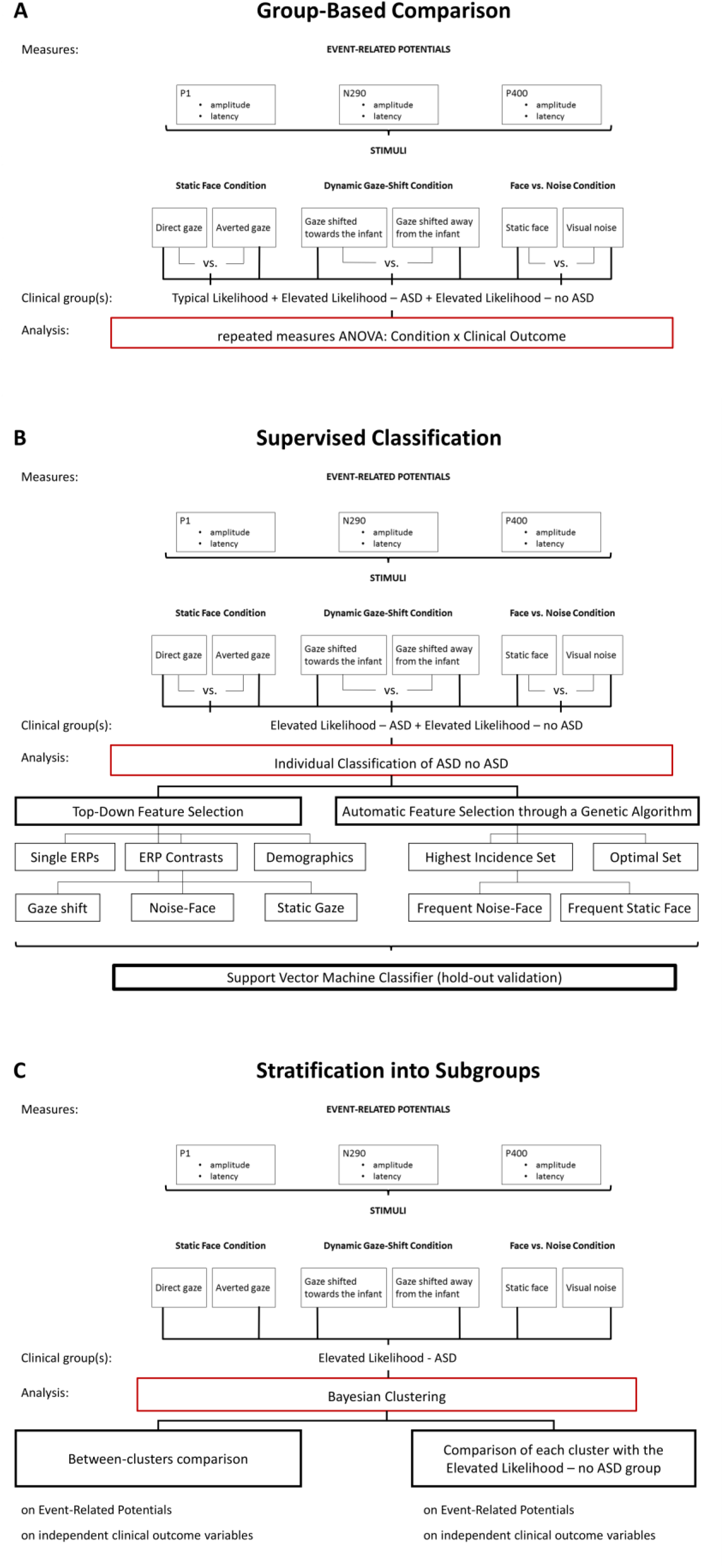
Flow chart of statistical analysis strategy.

#### Supervised Classification

Supervised classification was performed on EL infants test classification accuracy of ERP measures to predict clinical outcome at 36 months at an individual level *(Figure 1.B)*. A subsample of 144 EL infants was included in this analysis based on having at least 70% of ERP data available. Imputation through expectation maximization was used to handle missing data, which showed a pattern of data Missing At Random. Standardized average and differential ERP responses to the different stimuli conditions (see *Figure 1.B)* were included as input features for the classifier. Gender and age were also included as features to take into account their confounding effect (see *Table S1)*.

To investigate whether a subset of ERP measures, based on stimuli conditions or stage of processing, had a higher predictive value compared to the entire set, we performed top-down and bottom-up feature selection and compared classification performance of the entire feature set (‘all’ set) with that of the selected subsets (see *Figure 1.B)*. For top-down feature selection, we manually selected 6 different sets of features: (1) differential ERP responses to all stimuli conditions (‘ERP contrasts’); (2) averaged ERP responses to all conditions (‘single ERPs’); (3) averaged ERP responses to the Face-vs-Noise condition (‘noise-face’); (4) averaged ERP responses to the static face condition (‘static face’); (5) averaged ERP responses to the dynamic gaze shift condition (‘gaze shift’); (6) age and gender (‘demographics’). For bottom-up feature selection, we used a genetic algorithm ((Johannesson et al., 2002; Snaedal et al., 2012); *Supplementary Material)* based on the optimization of the Area Under the Curve (AUC) as an effective and combined measure of sensitivity and specificity, which allows to test the inherent ability of the predictor, providing a useful metric to evaluate diffusivity of the predictive features within the examined population (Kumar & Indrayan, 2011).

For classification, we used SVM classifiers with linear kernel. The sample was split into a main sample (70% of sample, n=101) for model selection, and a separate holdout sample (30% of sample, n=43) for validation. The sample partition was stratified for binary outcome (i.e., ASD vs. no-ASD). Each classifier was then fully cross-validated via 10-fold cross-validation on the main sample, with sample partitioning into folds stratified for binary outcome. The number of features was selected based on the AUC level reached during the evolutionary process, and stability of the process assessed through visual inspection. Once selected the number of features (n=21), the evolutionary process was repeated n=100 times to investigate the variability in the feature space. Feature sets with highest AUC (>85%) were used as input to a frequency analysis on the selected features, with incidence as an estimate of the relevance of each feature for the classification problem. The feature set providing the highest AUC (‘optimal’ set), the set of features with highest incidence (>80%, ‘highest incidence’ set), and condition-specific subsets of the ‘highest incidence set’ (‘frequent noise-face’ set, ‘frequent static face’ set) were selected as input for subsequent classifier analysis.

After feature selection, we performed classification through linear-kernel SVM classifiers built on the entire feature set and subsets of feature selection (see *Figure 1.B)*, trained on the main sample, and tested on the holdout validation sample. To evaluate classification performance, we computed AUC, sensitivity, specificity, accuracy, negative predictive power (NPV), and positive predictive power (PPV) from the ROC curve. 95% confidence intervals (CI) for each performance metric were computed using bootstrap with n=10000 repetitions. We tested for significant differences of classification performance, indexed by the AUC, with chance level prediction through a shuffle test with n=10000 repetitions (Golland & Fischl, 2003). The same procedure was used to test for significant differences in performance between the best performing classifier and the other classifiers. Analyses were completed using the *LIBSVM* toolbox (Chang, 2011) and custom scripts implemented on Matlab R2016b (MATLAB 9.1, The MathWorks Inc., Natick, MA, 2016).

#### Stratification into subgroups

To test whether neural processing of faces can define meaningful subgroups in ASD, we performed a clustering analysis within the EL-ASD group. We used Bayesian hierarchical clustering (BHC, (Savage et al., 2009)) on averaged ERP responses from infants in the EL-ASD group (n=32) to each condition (36 features, see Figure 1.C). BHC is a model-based clustering algorithm built on a Dirichlet process mixture to model uncertainty in the data. Compared to other clustering techniques, it overcomes common limitations such as relatively arbitrary selection of number of clusters, or distance metric (Marrelec, Messe, & Bellec, 2015). It uses, in fact, marginal likelihoods to decide which clusters to merge at each step of a bottom-up hierarchical clustering process. The use of Bayesian hypothesis-testing as model-based criterion for merging clusters is also advantageous in terms of quality of the resulting clusters compared to the use of ad-hoc distance metrics. As number of clusters is determined automatically in BHC, we tested stability of results through leave-one out cross-validation. Analysis were implemented using the *bhc* function from the *Bioconductor* package in *R (Savage R, 2019)*.

#### Comparison between clusters

To characterize the identified clusters, we investigated differences between clusters in face processing through t-tests. Next, we evaluated clustering performance by examining the association of cluster membership with clinical outcome variables at 36 months through t-tests. Specifically, we tested differences between clusters on symptoms, indexed by the ADOS Calibrated Severity Score (CSS) obtained from the raw total scores (CSS-Tot), Social Affect (CSS-SA) and Restricted and Repetitive Behaviors (CSS-RRB) domains; the ADI-R domain scores for the Social (ADI-Soc), Communication (ADI-Comm) and Repetitive Behaviours and Interests domains (ADI-RBI); and the SCQ total score (SCQ-Tot). Furthermore, we tested differences in developmental level, indexed by domain T-scores of MSEL: receptive language (RL), expressive language (EL), fine motor skills (FM) and visual receptive skills (VR). Finally, we tested differences in adaptive functioning, indexed by domain standard scores of VABS: communication, socialization, motor skills and daily living skills. Comparisons were implemented in *R*. Holm-Bonferroni correction of significance threshold (α) was used to correct significance of t-tests for multiple comparisons *separately* for the comparison between clusters on ERPs and on clinical outcome variables.

#### Comparison between clusters and EL-no ASD group

To understand if the diffuse pattern of face processing alterations identified as predictive of ASD by the supervised classification analysis was determined by subgroups of EL infants developing ASD having different face processing alteration profiles relative to the EL-no ASD group, we compared each cluster to the EL-no ASD group on ERP responses at 8 months. Comparisons were implemented in *R*, with Holm-Bonferroni correction for multiple comparisons.

## Results

### Group-level differences

We report group-level differences only when statistically significant *(Table S5)*.

We found a significant condition x outcome interaction on N290 latency in the face-noise contrast. Specifically, the EL-ASD group did not show a stimulus differentiation, while the TL (p=.010, d=0.61) and EL-no ASD (p=.021, d=0.52) groups showed longer latency to faces compared to noise, with no difference between TL and EL-no ASD groups (p=.551, d=0.10; *Figure* 2). This did not vary by cohort (F(1,170)=0.76, p=.386) and neither of the covariates had a significant interaction (ps>.41).

**Figure 2:**
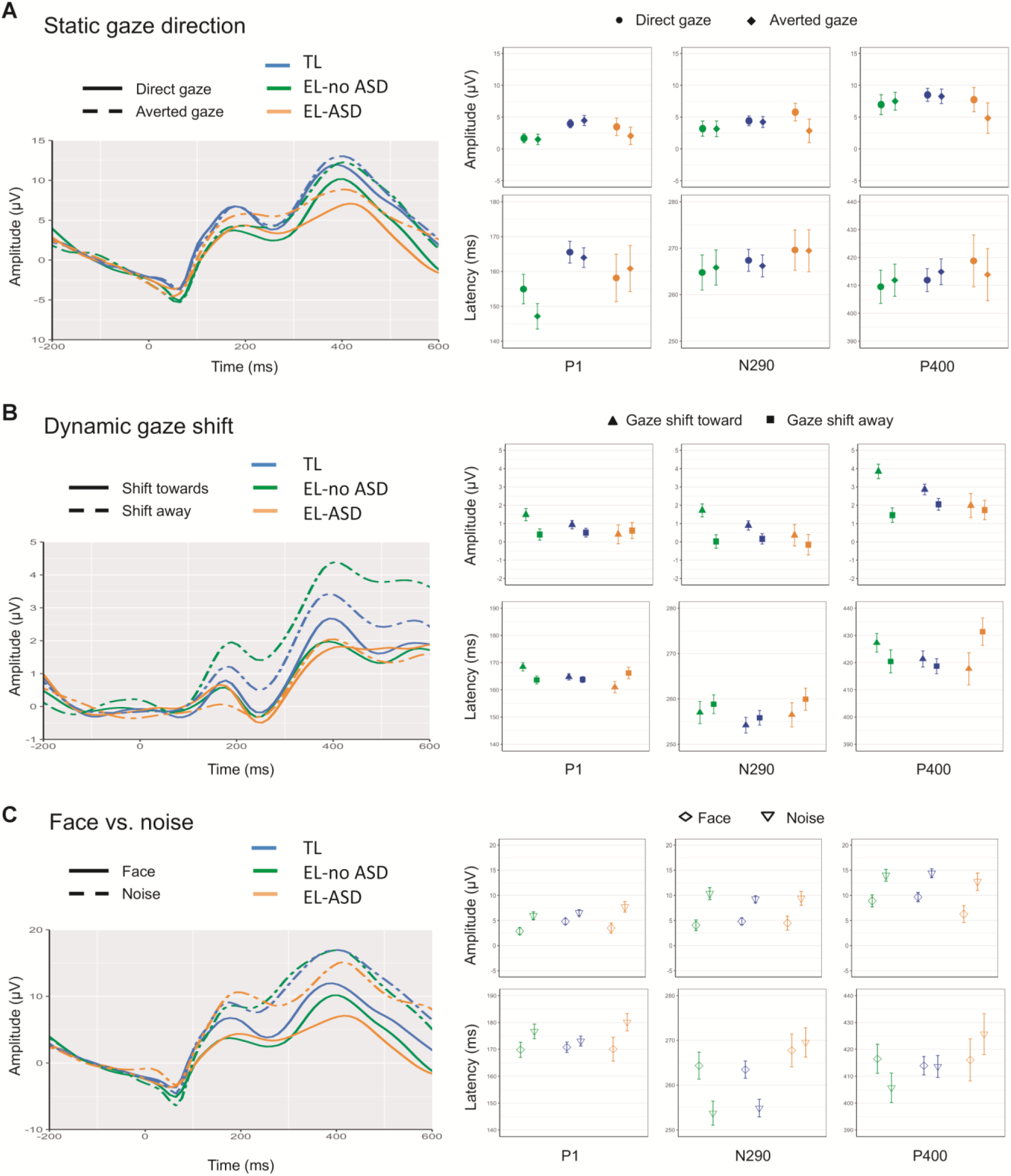
Grand average event-related potentials. Grand average ERPs across contrasts and groups over task-sensitive occipito-temporal channels (left) and means and standard errors of amplitude and latency of the three face-sensitive ERP components in each group (right).

There was a significant condition x outcome group interaction on P1 latency (F (2, 213)=4.95, p=.008), P400 amplitude (F (2,212)=4.13, p=.017) and latency (F(2, 208)= 3.51, p=.032). Specifically, the EL-ASD group had longer P1 latency to gaze shifting away versus towards, while the opposite effect was observed in the TL (p=.002, d=0.74) and EL-no ASD groups (p=.047, d=0.43), with no difference between TL and EL-no ASD (p=.122, d=0.26). There was no significant interaction with cohort (F (2,212)=1.48, p=.226), but the condition x outcome interaction became a trend when age and non-verbal ability were entered as covariates (F(2,201) = 2.45, p=.089). Lower non-verbal ability was associated with longer P1 latency to gaze shifting towards versus away (r=-.18, p=.008), with no association with age (r=.07, p=.32). Next, the EL-ASD group showed longer P400 latency to gaze shifting towards versus away from the viewer, with an opposite effect in TL (p=.011, d=0.55) and EL-no ASD (p=.021, d=0.47), and no significant difference between TL and EL-no ASD (p=.572, d=0.10). This did not vary by cohort (F(2,207)=0.91, p=.342) and was not influenced by covariates (ps>.24). Post-hoc t-tests revealed significant differences between EL-ASD and TL (p=.009, d=0.60) and EL-no ASD (p=.019, d=0.47), but not between TL and EL-no ASD (p=.488, d=0.12). See *Supplementary Material* for an additional analysis on the association between findings for P1 and P400 latency. Finally, there was enhanced P400 amplitude to gaze shifting towards versus away in the EL-ASD group (p=.016, d=0.46) and EL-no ASD group (p=.014, d=0.41), with no difference between EL groups (p=.482, d=0.12) and an opposite effect in the TL group. There was no interaction with cohort (F (1,211) = 0.27, p=.605) nor with covariates (ps>.19).

### Individual-level prediction

Combined sets of brain responses to different conditions of face stimuli provided the best predictive accuracy for ASD at an individual level. These combined sets provided, in fact, a predictive accuracy significantly different from chance level (‘ERP contrasts’, ‘Optimal’ and ‘Highest Incidence’ classifiers; see *Table S6/**Figure* 3), while classification performance for condition-specific subsets of features showed largely poor predictive power (see ‘Noise-Face’, ‘Static Face’, ‘Gaze Shift’, ‘Frequent Noise-Face’ and ‘Frequent Static Face’ classifiers in *Table S6)*. Specifically, the ‘Optimal’ set provided the best predictive algorithm for ASD outcome at 36 months, with an AUC of 77.1% (95% CI: [61.1, 90.5], p=0.01), significantly higher than classifiers built on condition-specific subsets selected top-down (see *Table S6)*. This suggests that automatic feature selection through the genetic algorithm outperforms top-down selection to indicate that combined sets of responses to different face stimuli improved predictive accuracy compared to condition-specific responses. Furthermore, compared to the ‘Highest Incidence’ set, which provided a nominally but not significantly higher classification accuracy (AUC=77.5%, 95% CI: [61.8, 90.2], p=0.02; p=0.15), the ‘Optimal’ set provided a better sensitivity to the ASD cases (see *Table S6)*. This suggests that, while the ‘Highest Incidence’ set indicates the most relevant features based on frequency of selection, there is additional useful information in the ‘Optimal’ set for prediction of ASD at an individual level (see *Table 1* for details on the features selected). Of note, sex was never selected by the genetic algorithm among sets providing the highest AUC, and age was selected less than 6% of the times, suggesting that the predictive model did not depend on the confounding variables. Details on classification performance from the different classifiers can be found in *Table S5*.

**Figure 3:**
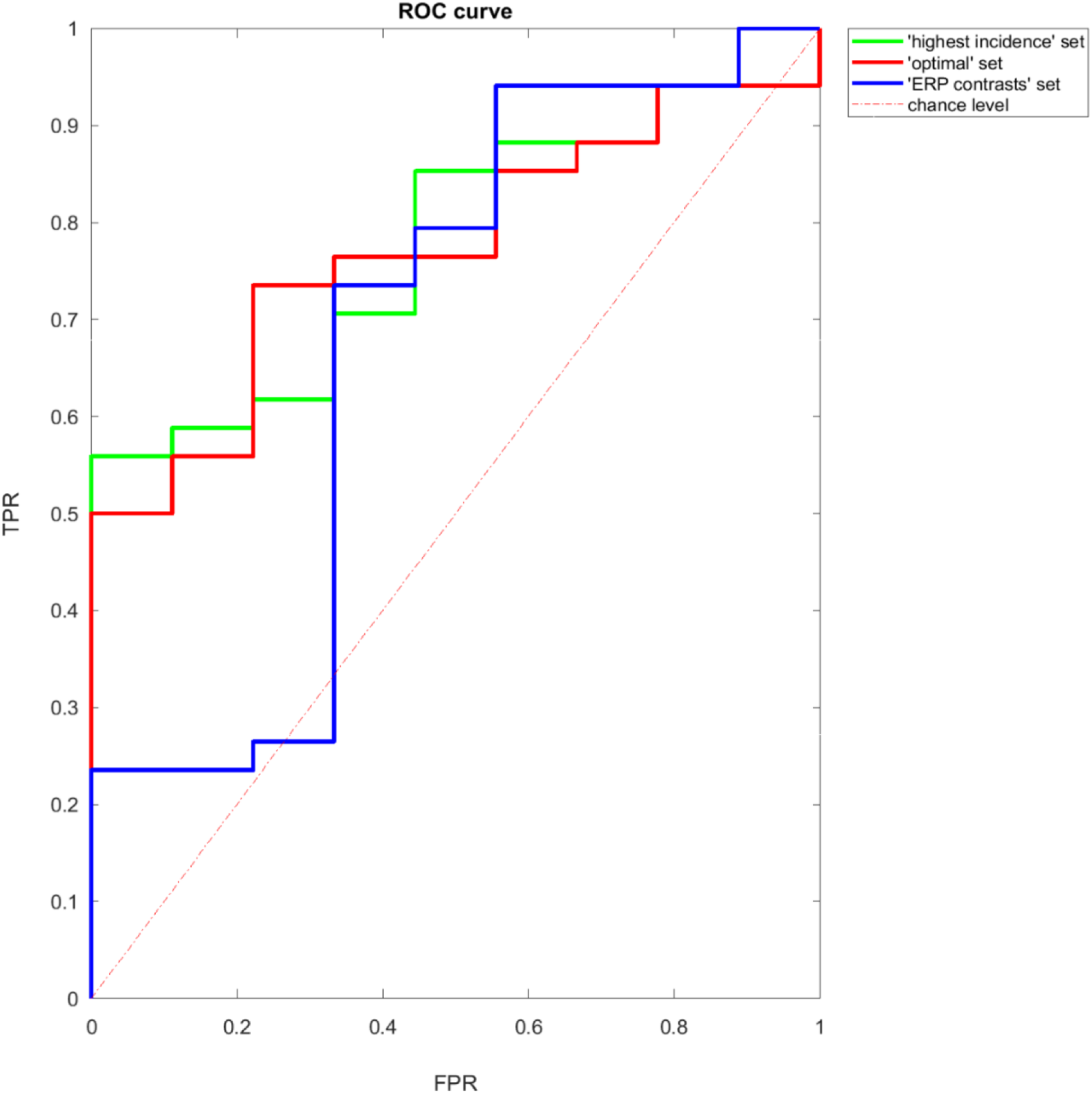
Classification performance. Receiver Operating Characteristic (ROC) curve for classifiers using different set of features to classify EL-ASD among EL siblings. Random predictors result in bisecting lines as ROC curves (red dashed line), while deviations in the upper hemifield indicate an increase in predictive accuracy. Only classifiers with a classification performance significantly different from chance level (assessed through a shuffle test) are included in this figure. *Abbreviations:* TPR = true positive rate or sensitivity; FPR = false positive rate, or 1-specificity.

**Table 1:**
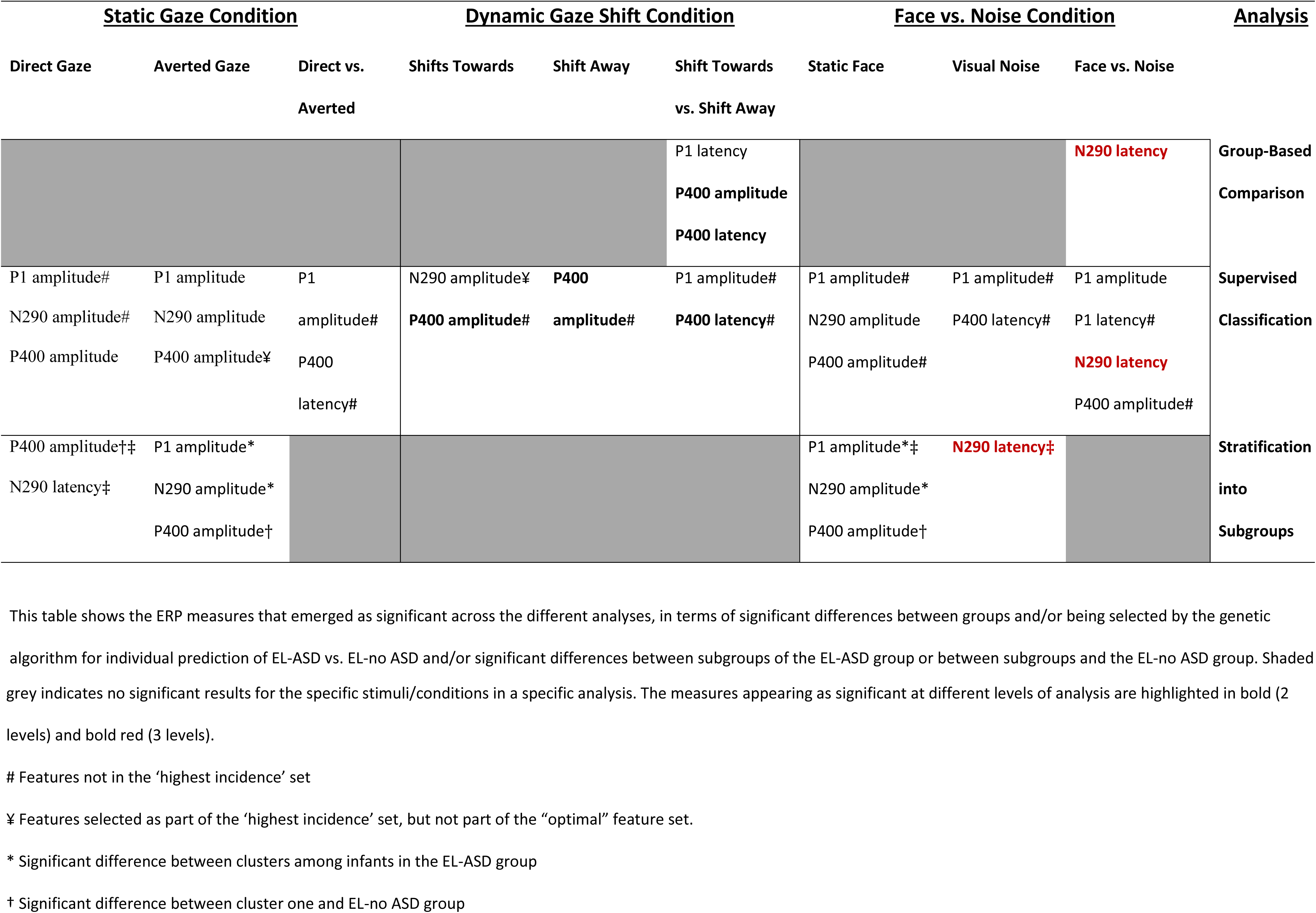
Summary of findings across different analyses

### Stratification into subgroups of ASD

The cluster analysis on average ERP responses to the different stimuli conditions (see *Figure 1.C)* identified two stable clusters among infants with later ASD diagnosis (*Figure* 4): cluster one consisted in 13 EL infants developing ASD (40.6% of the EL-ASD sample); cluster two consisted in 19 EL infants developing ASD (59.4%). Descriptive statistics for the different clusters are shown in *Table S7*.

**Figure 4.**
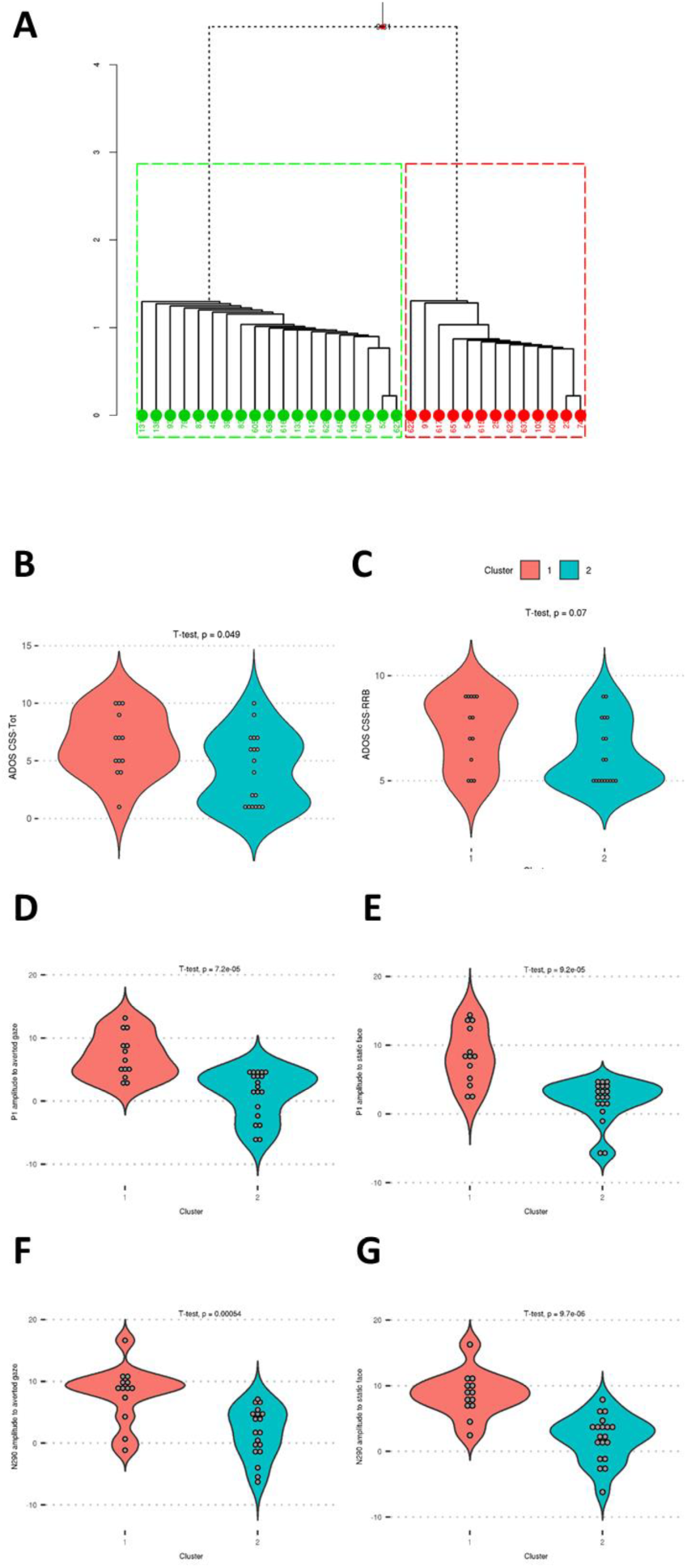
Stratification into subgroups of ASD based on face processing in infancy. Two clusters were identified among infants who meet diagnostic criteria for ASD at 36 months (A). Differences between clusters are shown in ASD symptoms severity (B), restricted repetitive behaviours (C), and ERP responses to face stimuli (D-to-G). Significance of the t-test for between-group differences is shown on each plot, not adjusted for multiple comparisons. *Abbreviations:* ERP = event-related potentials; ASD = autism spectrum disorder; ADOS = autism diagnostic observation schedule; CSS-Tot = total composite score from the ADOS; CSS-RRB = composite score for the restricted repetitive behavior subscale of the ADOS.

#### Comparison between clusters

Compared to cluster two, cluster one was characterized by higher P1 amplitude (t(21)=4.83, p<0.001) and N290 amplitude to static face, regardless of gaze direction (t(27)=5.45, p<0.001), and to faces with averted gaze (respectively t(27)=4.67 and t(24)=3.99, *ps*<0.001). Furthermore, infants from cluster one had significantly higher levels of ASD symptoms (ADOS CSS-Tot: t(27)=2.06, p=0.049), and marginally higher levels of restricted repetitive behaviours (ADOS CSS-RRB: p=0.07). However, differences were not significant after correcting for multiple comparisons (*Figure* 4).

#### Comparison between clusters and EL-no ASD group

Differences on face processing between ASD subgroups and EL infants who did not develop ASD were not significant after Holm-Bonferroni correction for multiple comparisons. Nevertheless, we found trend-level differences between groups indicating different alterations in neural processing of faces in infants from different ASD subgroups compared to siblings who did not develop ASD. In particular, compared to the EL-no ASD group, cluster one had reduced P400 amplitude in response to faces with direct gaze (unadjusted p=0.036), faces with averted gaze (unadjusted p=0.038), and irrespective of gaze direction (unadjusted p=0.049). Cluster two had, instead, reduced P1 amplitude to faces, irrespective of gaze direction (unadjusted p=0.032), reduced P400 amplitude to faces with direct gaze (unadjusted p=0.036), and longer N290 latency to faces with direct gaze (unadjusted p=0.026) and to visual noise (unadjusted p=0.006).

## Discussion

We present a set of analytic approaches to investigate *whether* and *how* a particular RDoC-defined domain is linked to a later diagnostic outcome (here ASD) in the context of a prospective design. Specifically, we focused on face processing as a putative precursor of the social features emerging later in development that are characteristic of ASD. Findings indicate alterations in both early sensory and later higher-level stages of neural processing of social (face/gaze) and scrambled face stimuli that differentiate infants with later ASD from those without later ASD across both group-based comparison, individual-level prediction and stratification *(Table* 1). These findings support a theoretical framework in which diffuse and individually heterogeneous anomalies in social and perceptual processing converge to contribute to the emergence of later categorical diagnosis of ASD.

### Neural processing of faces is diffusely atypical in ASD

Our analyses converged to support the contention that face processing is altered in early ASD. Specifically, the socially relevant processes of detecting a face and a shift in gaze are altered across a long time-course of information processing from the shortest latency components, and across multiple stimuli conditions *(Table* 1). N290 latency to visual noise versus faces emerged as a relevant feature across all three levels of analysis *(Table* 1), while P400 amplitude and latency to dynamic gaze shifts appeared to be informative features for prediction of ASD outcome at the level of groups and of individuals. The group of infants with later ASD (EL-ASD) showed a longer P400 latency and smaller P400 amplitude to gaze shifting towards versus away from the viewer and diminished effects of longer N290 latency to faces compared to visual noise compared to TL and EL-no-ASD groups. The same measures were part of a broader pattern predicting individual ASD diagnosis among siblings at elevated likelihood with approximately 77% accuracy. A broad pattern of alterations across the time-course of neural processing contributed to prediction at the individual level, including both shorter and longer latency components in response to different face conditions. Neural responses to dynamic gaze shifts alone, which appeared to be the best candidate precursors of ASD at a group level, did not provide sufficient predictive value on their own at an individual level *(Table S6)*. Despite significant differences between groups, there is a significant overlap in individual variation, suggesting that alterations in neural processing of dynamic gaze are neither necessary nor sufficient conditions for ASD development. Taken together, these findings support hypothesis (a) on diversity of altered face processing, which then likely reflects consistent differences in different specific aspects of face processing that are masked in different studies by their theory-driven focus on one or two components.

Our findings converge with a range of previous work in indicating the importance of early social processing and attention in the emergence of ASD. For example, a previous study of a subset of this cohort has showed that reduced P400 differentiation of dynamic gaze predicts ASD outcome in middle childhood, both in terms of stable diagnosis from 3 to 7 years and a ‘late’ diagnosis made after 3 years of age (Bedford et al., 2017). Furthermore, the reduced N290 latency difference between the social versus non-social stimuli in EL-ASD is in line with previous studies of EL siblings (E. Jones et al., 2016), and in the comparison of familiar and unfamiliar faces (Geraldine Dawson et al., 2002; Key & Stone, 2012). The N290/P400 complex is thought to be a developmental precursor to the N170 (de Haan, Johnson, & Halit, 2003), an established marker for social functioning (Neuhaus, Kresse, Faja, Bernier, & Webb, 2016) with a long history of research for alterations in ASD (Kang et al., 2018; McPartland, Dawson, Webb, Panagiotides, & Carver, 2004). Early-stage differences in neural processing, as we found here, may subsequently trigger a cascade of events that result in symptoms characteristic of ASD (Johnson, Gliga, Jones, & Charman, 2015). Reduced depth of processing for social stimuli may result, in fact, in failure to develop expertise in processing faces along a cumulative risk pathway.

### Stratification within ASD

The supervised classification analysis identified a diffuse pattern of neural responses to different face/noise stimuli at 8 months as predictive of individual ASD outcome in toddlerhood. To determine whether this pattern was determined by each infant with later ASD diagnosis having a different alteration in face processing at 8 months or by each infant having the same diffuse pattern of alterations, we performed a clustering analysis on ERP responses to the different conditions at 8 months among EL siblings developing ASD in toddlerhood *(Figure 1.C)*. We identified two different subgroups in ASD differing in intensity of response in early sensory (P1) and later high-order stages (N290) of processing faces with averted gaze, and static faces in general. Of note, differences between clusters in clinical outcome variables were not retained when correcting for multiple comparisons. While on the one hand it can be explained by a lack of statistical power, which might improve with increased sample size, this can be interpreted in light of a conceptualization of ASD as an epiphenomenon of earlier-interacting susceptibilities (Constantino, 2018). Although altered face processing represents one of these early measurable liabilities to ASD, it might not fully capture alone the heterogeneity in clinical expression of the disorder. It is also possible that the different clusters were defined by different patterns of interactions between ERP features, which was uncovered by the data-driven clustering approach, but could not be picked up by traditional group comparisons.

When compared to the broader group of infants who did not develop ASD, Cluster 1 showed reduced engagement to static faces (lower P400 amplitude), irrespective to gaze direction. This was associated with higher levels of ASD symptom severity, of restricted and repetitive behaviours, and of social-communication symptoms than their non-ASD peers, and also reduced motor, communication and daily living functioning skills *(Analysis S3)*. Cluster 2, instead, showed reduced perceptual capture to faces (smaller P1 amplitude), in particular to faces with averted gaze; reduced engagement to faces with direct gaze (lower P400 amplitude); and slower processing of faces with direct gaze and visual noise (longer N290 latency) compared to their non-ASD peers. This was associated with higher levels of ASD symptom severity, of restricted and repetitive behaviours, and of social-communication symptoms *(Analysis S3)*. Comparisons between ASD clusters and the EL-no ASD group on face processing suggest, therefore, that there may be some divergence within the ASD group. Thus, the diffuse pattern of features identified by the genetic algorithm as predictive of individual ASD outcome likely emerges from different subgroups in ASD being predicted by different features. Specifically, Cluster 1 appeared to be predicted by more specialized alterations in later stages of neural processing of static faces, while the other siblings developing ASD were predicted by a pattern of alterations in face processing that is more diffuse in terms of temporal stages, processing features (speed and/or intensity), and stimuli. Future work should integrate longitudinal measures to further refine our understanding of subgroups of face processing within ASD over development. Furthermore, future work might employ a different data-driven approach to parse individual heterogeneity of ASD and identify more homogeneous and replicable subgroups, which would potentially improve predictive accuracy and allow the identification of more specific physiological mechanisms (Bussu et al., 2019; Loth et al., 2017).

### Individual-level prediction: towards clinical utility?

The combination of ERP measures that best predicted ASD outcome at the individual level included responses to face/gaze and visual noise *(Table* 1), extending recent work using nonlinear features of EEG signals to support widespread dysfunction not specific to social processing (Bosl, Tager-Flusberg, & Nelson, 2018). Early intervention for infants at elevated likelihood for ASD, prior to the emergence of core ASD features, is supported and conducted at the group level (Green et al., 2015; Green et al., 2017; S. J. Webb, Jones, Kelly, & Dawson, 2014). Yet, individual prediction of ASD in the first year of life might be crucial to enable targeted intervention within a critical developmental window. We significantly reduced the age of detection compared to previous classification studies on behavioural measures (Bussu et al., 2018; Chawarska et al., 2014) and ERP parameters (Eldridge, Lane, Belkin, & Dennis, 2014). Individual-level prediction of ASD in the first year of life has shown more than 94% accuracy using functional and structural magnetic resonance imaging (Emerson et al., 2017; Hazlett et al., 2017). However, our results extend these findings by using ERPs to predict a more stable ASD diagnosis at 36 months in a larger EL sample. Furthermore, ERPs represent a more cost-effective, mobile, and infant-friendly neuroimaging technology, providing potential utility for inclusion as proxy outcome markers for intervention trials. To test this, future work should determine whether these parameters are sensitive to the effects of early intervention (E. J. H. Jones, Dawson, Kelly, Estes, & Jane Webb, 2017), together with tests of integration with other risk markers (e.g. genetic factors, brain imaging, parent-child interaction and behavioural measures) to improve individual prediction of ASD outcome. It is important to consider methodological limitations in machine learning related to relatively small sample size and model reliability (Yahata et al., 2017). Currently, there is no good theoretical justification for features selected, and results may vary accordingly. Although we validated our results on a separate, holdout sample, generalizability of the identified model must be tested through replication on an independent sample.

## Conclusion

Our three-pronged approach represents the first attempt to investigate robustness and generalizability of findings on a specific domain (social cognition/face processing) across different levels of analysis, from group-based comparisons to individual-level prediction of outcome and stratification. We focused on a domain in detail to show the analytical approaches necessary to investigate *whether* and *how* a specific domain contributes to the clinical category of ASD over development. Our findings show a diffuse pattern of atypicalities across the timecourse of neural processing in response to face and noise stimuli, suggesting that reported alterations likely reflect a more diffuse dysfunction that broadly affects neural processing of faces. These pattern of diffuse alterations was shared in good proportion across infants developing ASD; however, our clustering approach provided emerging evidence for subgroups of infants developing ASD that share more consistency in face processing. This adds to the literature illustrating early structural atypicalities (Hazlett et al., 2017) by showing early diffuse functional atypicalities, in line with the idea of ASD as a syndrome emerging from diffuse and interacting liabilities (Constantino, 2018). Using traditional and novel analytic approaches, we examined in detail the social cognition domain in infancy and investigated how this relates to the clinical ASD category in toddlerhood. Doing this, we addressed the emerging need in developmental neuroscience to incorporate RDoC constructs to improve our understanding of the mechanisms underlying ASD development.

## Data Availability

The datasets analysed during the current study are subject to the BASIS data sharing policy (http://www.basisnetwork.org/index.php?option=com_content&task=view&id=41&Itemid=68).

## Notes

Author Note Charlotte Tye, PhD, Department of Child & Adolescent Psychiatry and MRC Social, Genetic & Developmental Psychiatry Centre, Institute of Psychiatry, Psychology & Neuroscience, King’s College London; Giorgia Bussu, Department of Cognitive Neuroscience, Donders Institute for Brain, Cognition and Behavior, Radboud University Medical Center; Teodora Gliga, Centre for Brain and Cognitive Development, Birkbeck College, University of London; Mayada Elsabbagh, Department of Psychiatry, McGill University; Greg Pasco, Department of Psychology, Institute of Psychiatry, Psychology & Neuroscience, King’s College London; Kristinn Johnsen, Mentis Cura, Reykjavík, Iceland; Tony Charman, Department of Psychology, Institute of Psychiatry, Psychology & Neuroscience, King’s College London; South London and Maudsley NHS Foundation Trust (SLaM); Emily J. H. Jones, Centre for Brain and Cognitive Development, Birkbeck College, University of London; Jan K. Buitelaar, Department of Cognitive Neuroscience, Donders Institute for Brain, Cognition and Behavior, Radboud University Medical Center; Karakter Child and Adolescent Psychiatry University Centre; Mark H. Johnson, Centre for Brain and Cognitive Development, Birkbeck College, University of London; Department of Psychology, University of Cambridge; the BASIS team is listed in alphabetical order as follows: Anna Blasi, Simon Baron-Cohen, Rachael Bedford, Patrick Bolton, Susie Chandler, Celeste Cheung, Kim Davies, Janice Fernandes, Isobel Gammer, Holly Garwood, Jeanne Giraud, Anna Gui, Kristelle Hudry, Michelle Lieu, Evelyne Mercure, Sarah Lloyd-Fox, Helen Maris, Louise O’Hara, Andrew Pickles, Helena Ribeiro, Erica Salomone, Leslie Tucker, Agnes Volein. This study complies with APA ethical standards in data treatment. All procedures were in agreement with ethical approval granted by the London Central NREC (approval codes 06/MRE02/73, 08/H0718/76), and one or both parents gave informed consent to participate in the study. The datasets analysed in this study are subject to the BASIS data sharing policy. Previous studies have reported on partially overlapping datasets (Bedford et al., 2017; Mayada Elsabbagh et al., 2012; Salomone et al., 2018). This study was presented as a poster at the 10^th^ Donders Discussions in Nijmegen, 2017; and as an oral presentation at INSAR in Rotterdam, 2018, and at ETADE Brainview conference in London, 2018. We wish to thank all the individuals and families who participated in this research. This work was supported by MRC Programme Grant no. G0701484 and MR/K021389/1, the BASIS funding consortium led by Autistica (www.basisnetwork.org), EU-AIMS (the Innovative Medicines Initiative joint undertaking grant agreement no. 115300, resources of which are composed of financial contributions from the European Union’s Seventh Framework Programme (FP7/2007-2013) and EFPIA companies’ in-kind contribution), and AIMS-2-TRIALS (the Innovative Medicines Initiative 2 joint undertaking grant agreement no. 777394). Charlotte Tye is supported by a Tuberous Sclerosis Association fellowship and the NIHR Maudsley Biomedical Research Centre (BRC) at the Institute of Psychiatry, Psychology & Neuroscience and South London and Maudsley NHS Foundation Trust. Giorgia Bussu has received funding from the Marie Sklodowska Curie Actions of the European Community’s Horizon 2020 Program under grant agreement n°642996 (Brainview). Charlotte Tye and Giorgia Bussu share first authorship of this paper, while Emily Jones, Mark Johnson and Jan Buitelaar share last authorship.

### Competing Interest Statement

The authors have declared no competing interest.

### Funding Statement

This work was
supported by MRC Programme Grant no. G0701484 and MR/K021389/1, the BASIS funding consortium led
by Autistica (www.basisnetwork.org), EU-AIMS (the Innovative Medicines Initiative joint undertaking grant
agreement no. 115300, resources of which are composed of financial contributions from the European
Union’s Seventh Framework Programme (FP7/2007-2013) and EFPIA companies’ in-kind contribution), and
AIMS-2-TRIALS (the Innovative Medicines Initiative 2 joint undertaking grant agreement no. 777394).
Charlotte Tye is supported by a Tuberous Sclerosis Association fellowship and the NIHR Maudsley
Biomedical Research Centre (BRC) at the Institute of Psychiatry, Psychology & Neuroscience and South
London and Maudsley NHS Foundation Trust. Giorgia Bussu has received funding from the Marie
Sklodowska Curie Actions of the European Community’s Horizon 2020 Program under grant agreement
n°642996 (Brainview).

